# Genome-wide association study of DXA-derived hip morphology identifies associations with 4 loci in Chinese populations

**DOI:** 10.1101/2024.01.25.24301766

**Authors:** Jiayi Zheng, Jieyu Ge, Benjamin G. Faber, Huandong Lin, Raja Ebsim, Claudia Lindner, Timothy Cootes, Jin Li, Jonathan H. Tobias, Xin Gao, Sijia Wang

## Abstract

**Objective:** To identify genetic factors associated with hip morphology in Chinese populations.

**Methods:** An 85-point Statistical Shape Model (SSM) was applied to extract hip shape modes (HSMs). Diameter of the femoral head (DFH), femoral neck width (FNW) and hip axis length (HAL) were obtained from SSM points using Python scripts. Genome-wide association study (GWAS) was conducted in the Shanghai Changfeng (SC) cohort (N=5,310) for each phenotype of DXA-derived hip morphology. Replication of GWAS was conducted in the Core cohort (N=917).

**Results:** GWAS identified a total of 331 SNPs in 14 loci that were associated with features of hip morphology in the SC cohort. 4 of 14 loci were replicated in the Core cohort: rs143383 (*GDF5*) associated with HAL (P = 9.4×10^−10^), rs11614913 (*MIR196A2)* associated with HSM9 (P = 2.8 ×10^−10^), rs35049516 (*SUPT3H*) associated with HSM4 (P = 4.3 ×10^−10^) and rs7761119 (*UST*) associated with HSM8 (P = 1.7×10^−8^). Of these, two loci were known to affect hip morphology, including rs143383 (*GDF5*) and rs35049516 (*SUPT3H*), whereas rs11614913 (*MIR196A2*) and rs7761119 (*UST*) were novel. There was also overlap with previous GWAS of HSM and other hip-based metrics.

**Conclusions:** In the largest East Asian ancestry hip shape GWAS to date we identified and replicated four loci associated with different aspects of hip morphology *(GDF5, MIR196A2, SUPT3H, UST).* Strong SNP-to-gene evidence was found. All four loci have previously been implicated in musculoskeletal development, however this is the first report that rs11614913 (*MIR196A2)* and rs7761119 (*UST*) are associated with hip morphology. Despite the small sample size, this study paves the way for trans-ancestry meta-analyses.

## Introduction

Hip morphology is considered to be an important risk factor for hip osteoarthritis (HOA) (1). Our recent study found a much lower prevalence of radiographic HOA (rHOA) in the Shanghai Changfeng (SC) cohort, a community-based Chinese study, when compared to white participants in UK Biobank (UKB), a community-based UK study (2). These results were consistent with previous studies in other East Asian populations (3, 4, 5). In addition, distinct differences in hip morphology were seen between SC and UKB participants. For example, there was a lower prevalence of cam morphology, a narrower femoral neck, and a larger femoral head relative to femoral neck in the East Asian participants (SC) compared with the white participants (UKB). It is unclear if these differences in hip morphology and rHOA between SC and UKB are related, and to what extent these result from genetic as opposed to environmental differences.

Hip morphology derived from imaging can be characterized by statistical shape modelling (SSM), a form of principal component analysis, which computes orthogonal hip shape modes (HSMs) (6, 7). Whereas SSM describes the overall shape, geometric parameters measure specific hip morphology, for example, femoral neck width (FNW) or hip axis length (HAL). Previously, SSM-derived hip morphology formed the basis of a genome wide association study (GWAS) (8), which identified eight loci associated with pathways involved in endochondral bone formation. In addition, GWAS has examined geometric parameters of hip shape such as alpha angle (a measure of cam morphology) and minimum joint space width, providing valuable insights into osteoarthritis pathogenesis (9, 10). These studies were based in European populations, limiting the conclusions that could be applied to other ancestry groups.

Using previously derived measures of hip shape in SC, this study aims to provide the first comprehensive GWAS of hip shape in an East Asian population, including both HSMs and hip geometric parameters, namely FNW, HAL and diameter of the femoral head (DFH). Subsequently, we aimed to understand the extent to which hip shape associated loci overlap with other hip-based metrics including hip bone size, HOA, hip fracture and femoral neck bone mineral density (FN BMD).

## Materials and methods

### Participants in discovery set

SC consists of residents recruited from the Changfeng community of the Putuo District in Shanghai (N=6595, aged ≥ 45 years) between June 2009 and December 2012 (11). The Shanghai Changfeng Study is organized and directed by The Fudan-Erasmus Research Institute of Medicine (FERIM), which is a joint venture between Zhong Shan Hospital of Fudan University in Shanghai and the Erasmus Medical Center, Rotterdam, The Netherlands. Ethics approval was granted by the ethics committee of Zhongshan Hospital affiliated to Fudan University (B2008-119(3)) and written informed consent was provided by all participants before participation. All participants answered a survey and the majority had a high-resolution iDXA (GE Lunar) scans of their left hip (N=6082). Quality control identified 144 images of low-quality, 6 repetitious scans, and 622 samples without genomic data. The remaining 5310 samples were included in the GWAS analysis.

### Participants in replication

Participants in the Core cohort have been recruited by Fudan University in Shanghai since September 2018. The samples we included in this study were recruited before August 2022 (N=916, aged 20 to 60 years). High-resolution iDXA (GE Lunar) scans of their left hip were taken. All samples included in this study passed quality control of low-quality left hip DXA scans or low-quality gene data. Written informed consent was provided before participation. The Core cohort was recruited by Human Phenome Data Center of Fudan University and is still in expansion.

### Phenotyping

#### Statistical hip shape model

The left hip was outlined in DXAs using 85 points placed by a machine-learning trained software (BoneFinder®, The University of Manchester), excluding osteophytes. The point annotations were reviewed and corrected where necessary. Hip shape size and rotation were standardized by Procrustes analysis after point placement. Principal components analysis was then used to build a statistical shape model (SSM) from all available images in UKB, producing a set of orthogonal modes of variation. Further analysis focused on the first ten hip shape modes (HSMs), which explain 86.3% of hip shape variance. Applying the SSM built in UKB and the existing shape modes, all available images in the SC cohort and Core cohort were analysed to get comparable mode scores. A more detailed description of the methods, including positions of the 85 points outlining the hip, is provided in our previous publication(12).

### Hip geometric parameters

Custom Python 3.0 scripts were developed and used to automatically derive FNW, HAL and DFH, as previously described (13, 14). In brief, FNW was defined as the shortest distance measured between the superior and inferior side of the femoral neck, with a line-segment approach used to automatically calculate the narrowest distance between the relevant points. DFH was defined as the distance across the spherical aspect of the femoral head. To estimate this, a circle of best fit was placed around the femoral head, with the diameter of the circle taken to represent the DFH in mm. HAL was defined as the distance from the base of the greater trochanter to the medial aspect of the femoral head, drawn through the centre of the circle of best fit (used to calculate DFH), in mm. All images with geometric parameter values beyond ±2 standard deviations (SDs) from the mean were reviewed manually.

### Genotyping, imputation and quality control

Genotyping, imputation and quality control (QC) were performed for the SC cohort as previously described (15). Genomic DNA of the participants were extracted from peripheral blood leukocytes using QIAGEN (Hilden, Germany) QIAamp DNA Mini Blood Kit and genotyped with Illumina (San Diego, CA, USA) Infinium BeadChip genotyping array (707,180 markers). QC was carried out using PLINK v1.90b4 (16) (i.e. minor allele frequency (MAF) > 1%, Genotype missingness < 2%, Hardy-Weinberg p > 1e-5). Samples that had a sex discrepancy between genotypic and reported sex, or had genotype hard call rate <95%, or deviated from the expected inbreeding coefficient (-0.2<F<0.2) or were duplicated samples were excluded. We performed pre-phasing and phasing using SHAPEIT v2r790(17) and imputation with IMPUTE2 v2.3.1(18) with the 1000 Genomes (1000G) phase 3 data as the reference. QC was carried out again and 8,393,320 variants with call rate >98%, Hardy-Weinberg equilibrium P >1e-5, MAF >1%, and INFO score >0.8 were included in the final analysis. All genomic positions were in reference to hg19/build 37.

### Genome-wide association study (GWAS)

Analyses of HSMs were adjusted for age, sex and the first 10 ancestry principal components in the SC cohort and Core cohort. Height wasn’t included as a covariate in HSM analyses as HSM scores were already size adjusted by Procrustes analysis. As height is highly correlated with the hip geometric parameters (19), analyses of hip geometric parameters were adjusted for age, sex, height and the first 10 ancestry principal components in the SC cohort and Core cohort. GWAS was performed using mixed linear models in fastGWA (20). Significant threshold for our GWAS were set at 5×10^-8^. Manhattan plots and QQ plots were generated in the R package qqman(21). The genomic inflation factor (λ) was calculated to check for p value inflation on QQ plots. Conditionally independent variants in each GWAS were identified by GCTA-COJO (22, 23) with default parameters (-cojo-p 5×10^-8^, -cojo-wind 10,000kb). Physically closest gene of variants were annotated by dbSNP (https://www.ncbi.nlm.nih.gov/SNP/). Regional linkage disequilibrium plots and association plots were generated by LocusZoom(24).

### SNP heritability and genetic correlation

SNP-based heritability was estimated using GCTA-GREML(22). Genetic correlations between hip morphology phenotypes and hip bone mineral density of both femoral neck and total hip were calculated by a bivariate GREML analysis in GCTA(25). Hip bone mineral density was measured by iDXA (GE Lunar) in the SC cohort.

### Overlap with GWAS of other traits

#### GWAS of hip-based metrics

All lead SNPs found in the SC hip morphology GWAS were looked up in GWAS of DXA bone area of the hip (26), FN BMD(27) and hip fracture(28), and also in the largest osteoarthritis (OA) GWAS (29).

#### GWAS of hip shape modes

Loci associated with HSMs in the previous European GWAS of HSMs (8) were looked up in our Chinese HSM GWAS.

### Fine mapping

Functional Mapping and Annotation (FUMA)(30) was applied to identify candidate genes for each lead variant. CADD v1.3 (Combined Annotation Dependent Depletion) were used to evaluate the functional consequences of the variants (31). Regulatory elements of non-coding human genome were identified using RegulomeDB(32). ChromoHMM were used to the prediction of chromatin state (core 15-state model) in chondrocytes and osteoblasts(33). Significant expression quantitative trait locus (eQTL) association (GTEx v8 database, false discovery rate <0.05) (34, 35, 36) with the lead variant were looked up.

## Results

### Hip morphology GWAS

In this study, we conducted a GWAS for 13 measures of hip morphology including 10 HSMs, FNW, HAL and DFH. Our discovery GWAS were conducted in the SC cohort, comprising 5310 individuals (3047 females and 2263 males) with an average age of 63.4 years (SD: 9.4 years; range: 46-96 years) (Supplementary Table 1). Replication took place in the Core cohort, consisting of 917 individuals (538 females and 379 males) with a mean age of 33.5 years (SD: 10.7 years; range: 20-60 years) (Supplementary Table 1). The Manhattan plot identified 14 genome-wide significant (P<5×10^-8^) independent loci in discovery GWAS for the first 10 hip shape modes and hip geometric parameters (Figure 1). Of these, 4 loci were replicated in the Core cohort with the same direction of effect and a P value < 0.05, including rs35049516 (*SUPT3H*) of HSM4, rs7761119 (*UST*) of HSM8, rs11614913 (*MIR196A2*) of HSM9, and rs143383 (*GDF5*) of HAL (Table 1). QQ plots showed no inflation of p values with all genomic inflation factors under 1.04 (Supplementary Figure 1 and Supplementary Table 4). The regional information generated by Locuszoom indicated the strength and extent of the association signal relative to genomic position (Figure 2, Supplementary Figure 2). Relatively high SNP heritability was estimated by GCTA in hip morphology GWAS (HSM6: *h*^2^=0.49 [95% CI 0.38, 0.59]; HSM3: 0.47 [0.36, 0.57]; HAL: 0.50 [0.40, 0.61]; FNW: 0.46 [0.35, 0.57]; DFH: 0.53 [0.42, 0.64]); HSM5 was estimated to have a small SNP heritable component (HSM5: 0.17 [0.07, 0.28]) (Supplementary Table 5). The bivariate GREML analysis showed moderate genetic correlation between HAL and FN BMD (rG: 0.46 [0.27, 0.66]), and also between HSM5 and total hip BMD (0.47 [0.14, 0.79]) (Supplementary Table 6).

**Figure 1:**
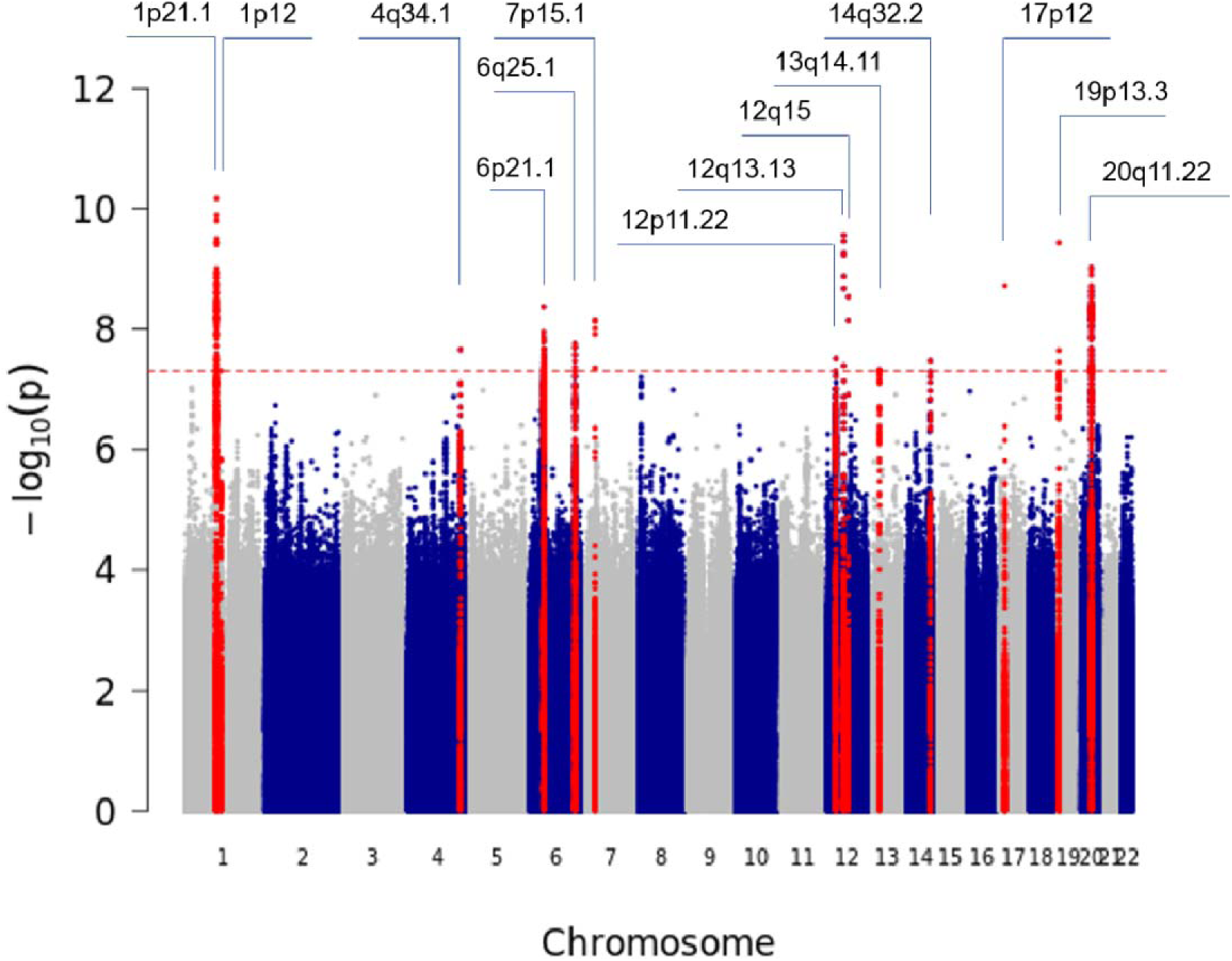
Manhattan plot of the discovery GWAS in the SC cohort.

**Figure 2:**
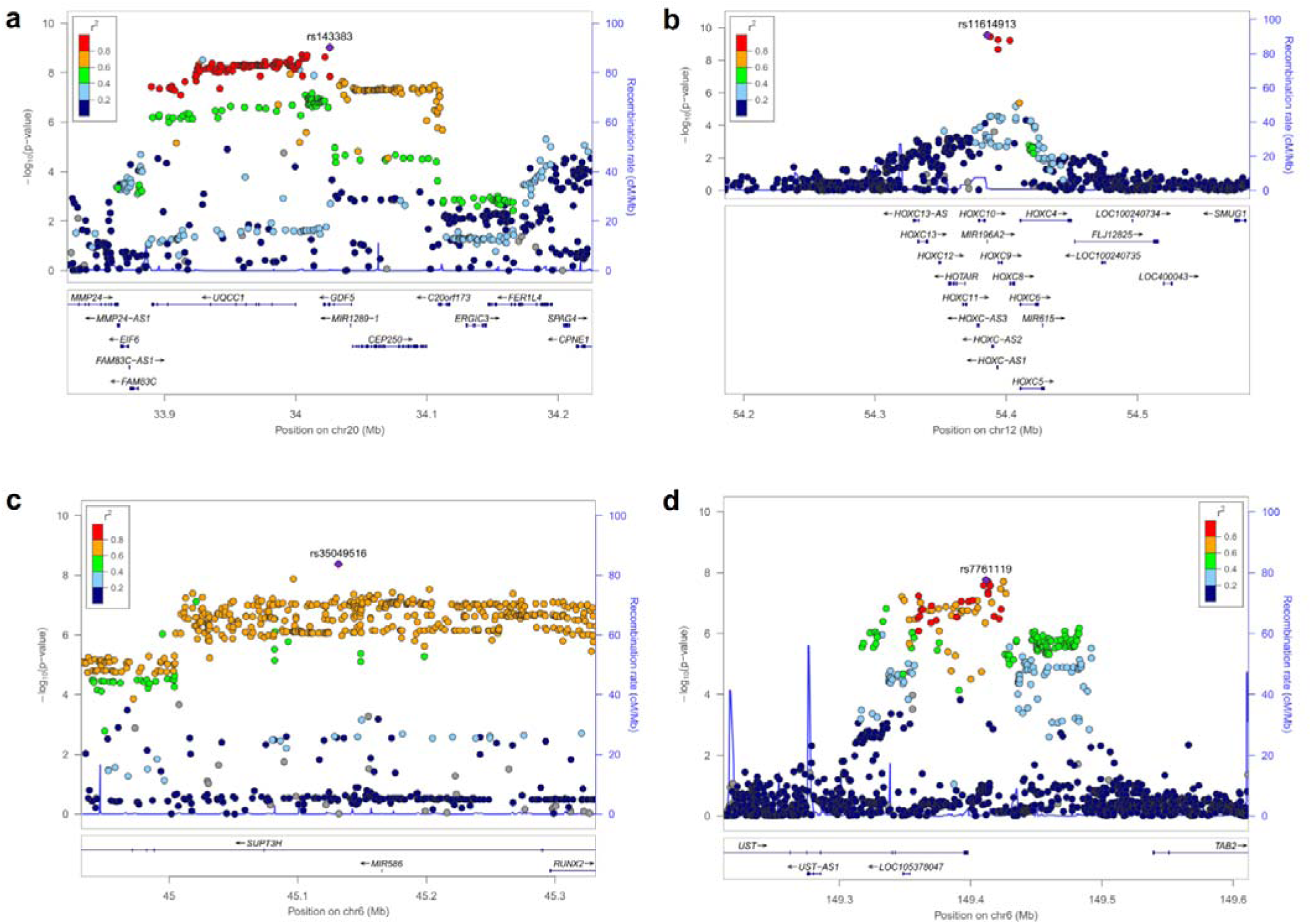
Locuszoom plots of the four replicated loci.

**Figure 3:**
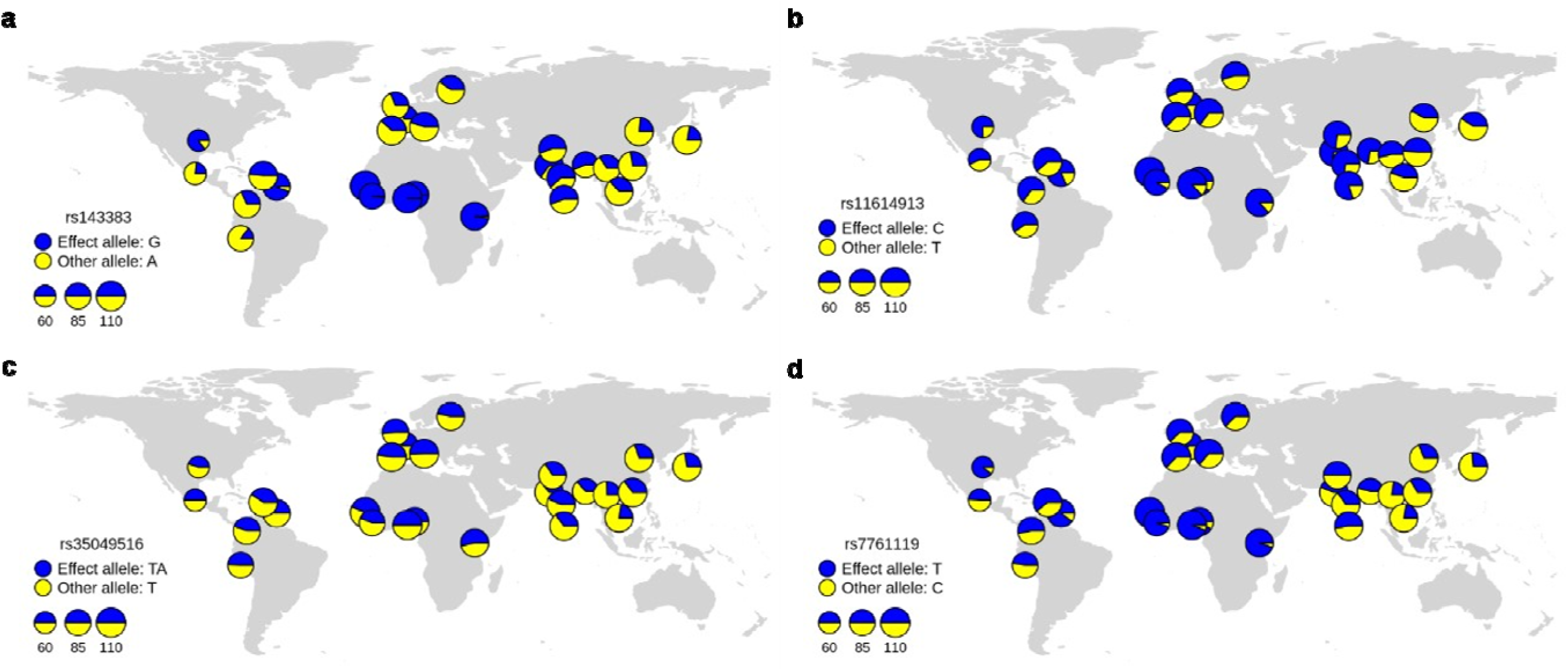
Geographical distribution of associated SNP allele frequencies. Allele frequencies (a) rs143383 (GDF5), (b) rs11614913 (MIR196A2), (C) rs35049516 (SUPT3H) and (d) rs7761119 (UST) were obtained from the 1000 Genome Project and visualized by the R package rworldmap. Effect alleles are marked by blue, and other alleles are marked by yellow. Each pie chart reflects the sample size and frequency of alleles in each subgroup.

**Table 1.**
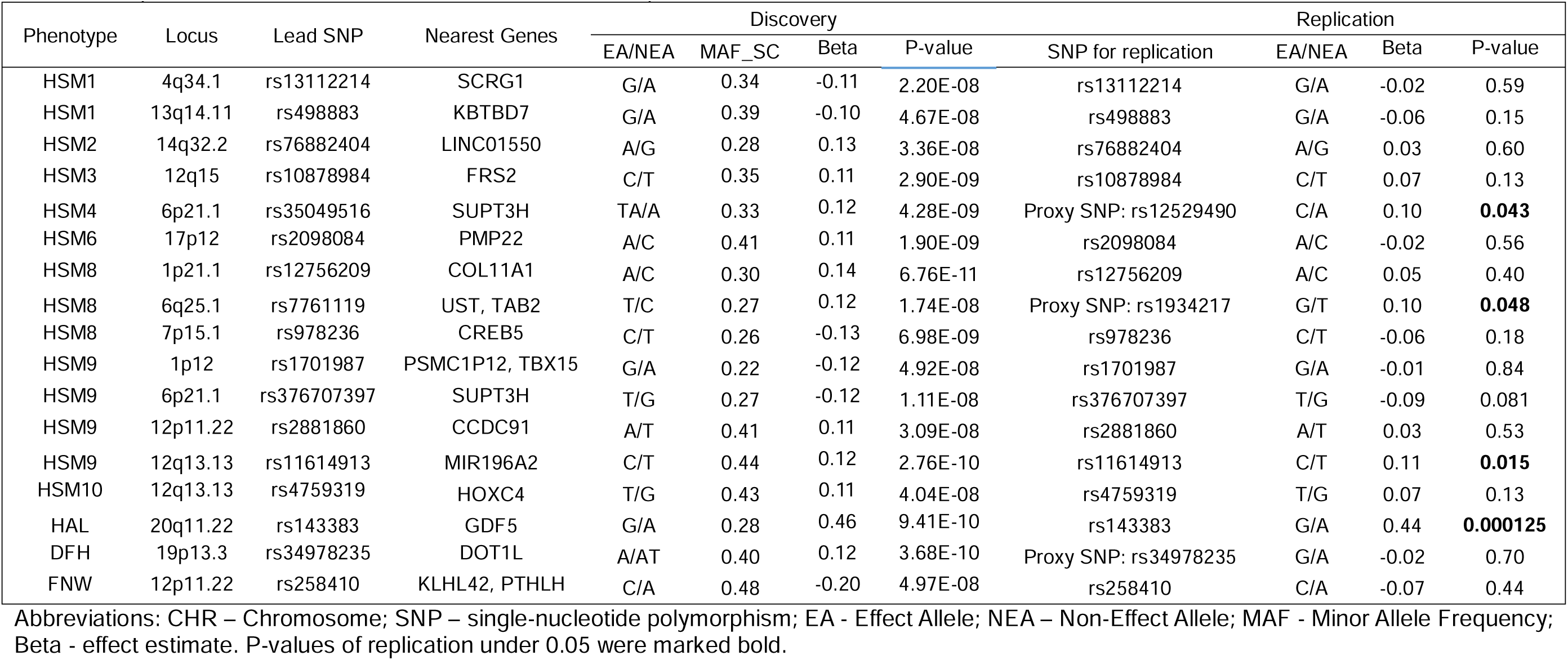
Independent loci identified in the SC cohort and the replication in the Core cohort.

**Table 2.** SNP associations with previous GWAS of OA and hip-based metrics.

| Shanghai Changfeng cohort |  |  |  |  | GO Consortium (All OA) |  | GO Consortium (HOA) |  | Hip fracture |  | FN BMD |  | DXA Bone Area (Total hip area) |  |
| --- | --- | --- | --- | --- | --- | --- | --- | --- | --- | --- | --- | --- | --- | --- |
| Pheno | Lead SNP | Locus | EA | Beta | Beta | P-value | Beta | P-value | Beta | P-value | Beta | P-value | Beta | P-value |
| HSM1 | rs13112214 | 4q34.1 | G | -0.11 | -4.00E-04 | 0.94 | -0.0067 | 0.52 | -0.0097 | 0.5515 | 0.0087 | 0.6339 | 0.013 | 0.12 |
| HSM1 | rs498883 | 13q14.11 | G | -0.10 | -0.0014 | 0.83 | 0.0153 | 0.23 | -0.0072 | 0.7166 | -0.0112 | 0.0556 | -0.032 | **7.21E-03** |
| HSM2 | rs76882404 | 14q32.2 | A | 0.13 | 0.01 | 0.31 | 0.0366 | 0.07 | -0.0553 | 0.08258 | 0.0158 | 0.7471 | -0.028 | 0.17 |
| HSM3 | rs10878984 | 12q15 | C | 0.11 | -0.017 | **4.68E-04** | -0.0311 | **9.20E-04** | -0.0409 | **0.007723** | -0.0080 | 0.0684 | -0.05 | **1.87E-09** |
| HSM4 | rs35049516 | 6p21.1 | TA | 0.12 | -0.016 | **6.06E-03** | -0.058 | **1.85E-05** | NA | NA | NA | NA | NA | NA |
| HSM6 | rs2098084 | 17p12 | A | 0.11 | 0.0053 | 0.52 | 0.0223 | 0.18 | 0.0044 | 0.8728 | 0.0152 | 0.8151 | 0.028 | **0.049** |
| HSM8 | rs12756209 | 1p21.1 | A | 0.14 | -0.022 | **1.19E-04** | -0.0666 | **3.87E-08** | 0.039 | **0.03083** | -0.0218 | 0.0568 | 0.079 | **4.45E-21** |
| HSM8 | rs7761119 | 6q25.1 | T | 0.12 | 0.007 | 0.13 | -0.0053 | 0.57 | -0.0084 | 0.5803 | 0.0013 | 0.8728 | -0.017 | **0.043** |
| HSM8 | rs978236 | 7p15.1 | C | -0.13 | -0.0036 | 0.43 | -0.0181 | **0.044** | -0.0016 | 0.9131 | 0.0112 | 0.1439 | 0.00053 | 0.95 |
| HSM9 | rs1701987 | 1p12 | G | -0.12 | -0.0037 | 0.41 | 0.0046 | 0.61 | -0.0214 | 0.1468 | -0.0080 | 0.3005 | 0.00034 | 0.97 |
| HSM9 | rs376707397 | 6p21.1 | T | -0.12 | -0.0132 | 0.14 | -0.0089 | 0.67 | -0.0101 | 0.7194 | NA | NA | NA | NA |
| HSM9 | rs2881860 | 12p11.22 | A | 0.11 | -0.0055 | 0.31 | -0.038 | **3.20E-04** | 0.0655 | **1.609E-04** | -0.0236 | **0.0102** | 0.029 | **3.42E-03** |
| HSM9 | rs11614913 | 12q13.13 | C | 0.12 | 0.0027 | 0.55 | 0.0065 | 0.48 | -0.0782 | **1.297E-07** | NA | NA | -0.0095 | 0.25 |
| HSM10 | rs4759319 | 12q13.13 | T | 0.11 | -0.0047 | 0.32 | -0.0028 | 0.76 | 0.0741 | **1.358E-06** | -0.0409 | **3.23E-07** | 0.0075 | 0.37 |
| HAL | rs143383 | 20q11.22 | G | 0.46 | -0.032 | **1.06E-11** | -0.0254 | **6.61E-03** | 0.0115 | 0.4459 | 0.0168 | 0.073586 | 0.08 | **1.71E-20** |
| DFH | rs34978235 | 19p13.3 | A | 0.12 | -0.0028 | 0.68 | NA | NA | NA | NA | NA | NA | NA | NA |
| FNW | rs258410 | 12p11.22 | C | -0.20 | 0.0123 | **7.67E-03** | 0.0576 | **2.73E-10** | -0.0378 | **0.01131** | -0.0096 | 0.2128 | -0.024 | **4.08E-03** |

Abbreviations : CHR – Chromosome; BP – Position in base pairs; EA – Effect Allele; Beta – Effect estimate; GO – Genetics of Osteoarthritis; HOA – Hip osteoarthritis; FN BMD – Femoral Neck Bone Mineral Density. P-values under 0.05 were marked bold.

### SNP functional annotation

Two of the four SNPs found in the SC cohort to be associated with hip morphology and successfully replicated in the Core Cohort had a high CADD score indicating these might be deleterious SNPs (Table 3). Rs11614913, an exonic SNP in *MIR196A2* (noncoding RNA) associated with HSM9, had a CADD score of 21.7 and a RegulomeDB score of 5. Moreover, rs143383, a SNP in UTR5 area of gene *GDF5*, had a CADD score of 21.2. Of those four replicated SNPs, three SNPs showed significant eQTL associations with genes in GTEx (Table 3). Rs143383 was found to be an eQTL for several genes with the strongest evidence being for *UQCC1* (P 2.9×10^-57^, tissue cultured fibroblasts) and *GDF5* (P 8.6×10^-15^, tissue esophagus). Rs7761119 was found to be an eQTL for *UST-AS2* (P 1.9×10^-7^, tissue cultured fibroblasts) and *TAB2* (P 1.2×10^-6^, tissue cultured fibroblasts). Rs11614913 was also found to be an eQTL in fibroblasts for *HOXC-AS1* (P 2.2×10^-12^), *GPR84* (P 6.6×10^-5^) and *HOXC8* (P 8.3×10^-5^) (all eQTLs information in supplementary table 2).

**Table 3.** SNP function of the replicated lead SNPs.

| Phenotype | Lead SNP | Locus | Func | CADD | RDB | Chromatin state<br>chondrocyte | Chromatin<br>state<br>osteoblast | eQTL |
| --- | --- | --- | --- | --- | --- | --- | --- | --- |
| HSM4 | rs35049516 | 6p21.1 | intronic | 0.552 | NA | 15 | 15 | NA |
| HSM8 | rs7761119 | 6q25.1 | intergenic | 0.091 | 4 | 15 | 15 | *UST-AS2*, *TAB2* in fibroblasts |
| HSM9 | rs11614913 | 12q13.13 | ncRNA_exoni<br>c | 21.7 | 5 | 14 | 7 | *HOXC-AS1*, *GPR84*, *HOXC8* in<br>fibroblasts |
| HAL | rs143383 | 20q11.22 | UTR5 | 21.2 | NA | 15 | 15 | *UQCC1* in fibroblasts, *GDF5* in<br>esophagus |

Abbreviations: SNP – single-nucleotide polymorphism; CHR – Chromosome; BP – Position in base pairs.
Func: Functional consequence of the SNP on the gene obtained from ANNOVAR.
CADD: CADD score which is computed based on 63 annotations.
RDB: RegulomeDB score which is the categorical score (from 1a to 7). 1a is the highest score that the SNP has the most biological evidence to be regulatory element.
Chromatin state chondrocyte (from 1 to 15, lower number higher activity).
Chromatin state osteoblast (from 1 to 15, lower number higher activity).

### Association with other bone related phenotypes

Associations with OA and other hip-based metrics were evaluated for the four SNPs found in the SC cohort and successfully replicated in the Core Cohort (Table 3). Rs35049516 (*SUPT3H*) was found to be protective of OA (β -0.016, P 6.06×10^-3^) and and HOA (β -0.058, P 1.85×10^-5^) and positively associated with HSM4 (a narrower femoral neck and larger femoral head, β 0.12, P 4.79×10^-9^). Rs143383 (*GDF5*) was found to be protective of both OA (β -0.03, P 1.06×10^-11^) and HOA (β -0.025, P 6.61×10^-3^) and positively associated with HAL (β 0.46, P 1.07×10^-9^). Rs11614913 (*MIR196A2*), which was positively associated with HSM9 (a wider femoral neck and larger femoral head, β 0.12, P 2.76×10^-10^) was protective of hip fracture (β -0.078, P 1.30×10^-7^).

### Overlap with previous largest HSM GWAS study

Nine SNPs previously found to be associated with a HSM in a European ancestry GWAS showed nominal significance in the SC GWAS (P<0.05)(8) (Supplementary Table 3). Seven of nine SNPs were replicated in the SC with a P value<0.05. For example, rs2158915 (*SOX9*) was significantly in negative association with HSM1 in Caucasian cohorts (β -0.13, P 8.47×10^-27^), which was also associated with HSM2 (β -0.06, P 0.03), HSM3 (β -0.07, P 0.0037) and HSM4 (β -0.05, P 0.05) in the SC cohort. Rs1243579, positively associated with HSM1 in the European ancestry GWAS (β 0.12, P 2.85×10^-14^), also showed strong association with HSM2 (β 0.11, P 5.06×10^-7^) and HSM3 (β 0.09, P 3.01×10^-6^) in the SC cohort.

## Discussion

In this study we present the results from a GWAS of 13 measures of hip morphology (ten orthogonal HSMs and three hip geometric parameters) that identified 17 conditionally independent SNPs in the SC cohort. Of these, 4 loci were replicated in an independent replication called the Core cohort (rs11614913 in *MIR196A2*, rs143383 in *GDF5*, rs35049516 in *SUPT3H* and rs7761119 at 6q25.1 near *UST* and *TAB2*). This is the first hip morphology GWAS performed in a Chinese population and replicated in an independent Chinese cohort. Although in European populations larger GWAS on HSMs, AA and minimum joint space width were carried out to decipher genetic architecture of hip shape and provide insights to osteoarthritis pathogenesis, this study provided genetic characteristics of non-European populations.

Recent observational studies found that cam morphology, larger lesser trochanter size and wider FNW are associated with higher prevalence of HOA (6, 37, 38, 39). In our study, we found evidence of overlapping associations between certain genetic loci and hip morphology and HOA (29). Of the 4 replicated loci, rs35049516 (*SUPT3H*) and rs143383 (*GDF5*), which were associated with HSM4 (a narrower femoral neck and larger femoral head) and larger HAL respectively, were both protective of OA and HOA. This indicates a shared genetic basis between hip morphology and HOA in East Asians, as previously found in European cohorts (8).

One of the replicated SNPs, rs11614913 located in the microRNA *MIR196A2* gene and the HOXC gene cluster of developmental transcription factors(40), was found to be an eQTL for *HOXC-AS1, GPR84, HOXC8* in cultured fibroblasts. Rs11614913 (*MIR196A2*) has been reported to be associated with bone size of lumbar spine area from DXA scans but has not previously been associated with hip morphology. Styrkarsdottir et al. performed a GWAS of bone size and found rs11614913 (*MIR196A2*) as the strongest signal. They suspected the variant might affect the efficiency of miR-196-5p target gene suppression. Then they conducted experimental induction of *MIR196A2* (C-alleles or T-alleles) into HEK293T cells and assessed the mRNA expression by RNA-sequencing. The gene enrichment for the entire set of repressed genes due to experimental induction of *MIR196A2* were reported to find embryonic skeletal system morphogenesis, supporting the relevance of *MIR196A2* for bone morphology (26).

The other locus first reported to be associated with hip morphology was rs7761119, an intergenic SNP located near *UST* and *TAB2*. This was found to be an eQTL for both *UST* and *TAB2* in cultured fibroblasts in GTEx. *TAB2*, TGF-Beta Activated Kinase 1 (MAP3K7) Binding Protein 2, is known to be involved in the osteoclast differentiation KEGG pathway, consistent with a role in hip morphology.

Rs35049516 was an intronic of *SUPT3H*, which was reported to be associated with body height and heel bone mineral density (41, 42). In a recent GWAS of minimum joint space width (mJSW) in European populations, *SUPT3H* was reported to associated with mJSW, an OA-related hip morphology (10). The overlap of findings indicated Chinese and Western populations appear to have similar genetic influences on hip morphology.

Rs143383 (*GDF5*), a functional SNP in the 5’-untranslated region of GDF5, was responsible not only for developmental dysplasia of the hip (DDH) but also lumbar disc degeneration and osteoarthritis (43) Miyamoto et al firstly reported that rs143383 showed significant association with hip and knee osteoarthritis in East Asian populations, suggesting that decreased GDF5 expression is involved in the pathogenesis of osteoarthritis (44). Dai et al. showed there was an association of rs143383 and DDH susceptibility in a Chinese Han population by a case-control study (45). Although GDF5 has been known to be associated with HAL in female of European ancestry (46), this is the first report of its association with HAL in East Asian populations. Wu et al found evidence that rs143384 and rs143383 A alleles both have high frequencies in non-Africans and show strong extended haplotype homozygosity and high population differentiation in East Asians, indicating that positive selection has driven the rapid evolution of these two variants of the human GDF5 gene (47). Besides, rs143383 (*GDF5*) was an eQTL for not only *GDF5* but *UQCC1*, which was an OA candidate gene and might be a relevant gene to hip morphology. In a GWAS study in the Chinese Han population, rs6060373(*UQCC1*) was shown to play a role in the etiology of DDH(48). In contrast, in a case-control study of a Turkish population, *UQCC1* rs6060373 polymorphisms were not associated with DDH(49). In a recent alpha angle (a proxy for cam morphology) meta GWAS in the European populations, *UQCC1* was found as a candidate gene for cam morphology(9). Our finding supported the sharing relevance of *UQCC1* of hip morphology in Chinese and European populations.

Our study has several strengths. Firstly, we conducted the first hip morphology GWAS in an East Asian population, broadening the ancestral diversity of GWAS looking at hip morphology. Secondly, advanced machine-learning algorithms were applied to hip DXA scans to extract hip shape modes and hip geometric parameters, which mirror those applied in UK Biobank. This provides opportunities for trans-ancestral hip morphology GWAS in the future. There are also some limitations to our study. The sample size is limited by the small number of East Asian studies that contain hip imaging and paired genetic data, which precluded the use of techniques such as genetic correlation or well-powered Mendelian randomization (50, 51). That said, we were sufficiently powered to discover and replicate 4 loci. Further work is needed to develop large biobanks containing genetic and imaging data in East Asian populations to allow for comparisons with predominately European ancestral studies such as UK Biobank and FinnGen (52, 53). The SC cohort is community-based but not multi-regional, so is not entirely representative of Chinese populations.

In conclusion, our GWAS of hip morphology in Chinese populations identified four loci, namely rs35049516 (*SUPT3H*) for HSM4, rs7761119 (*UST*) for HSM8, rs11614913 (*MIR196A2)* for HSM9 and rs143383 (GDF5) for HAL. These have previously been suggested to play an important role in the musculoskeletal system. However, the present study provides the first evidence that they also play a similar role in Chinese populations. Further, we identify two novel independent significant loci, rs11614913 (*MIR196A2)* and rs7761119 (*UST*), which have not previously been associated with hip morphology. Future studies with larger sample size and multi-centered Chinese cohorts might be needed to decipher the genetic factors of hip morphology in the Chinese population.

## Data Availability

All data produced in the present study are available upon reasonable request to the authors

## Acknowledgement

We express our deep appreciation to the members of the Department of Endocrinology and Metabolism, Zhongshan Hospital for their assistance for collecting data of the SC cohort. Our study is also supported by the Human Phenome Data Center of Fudan University. SW is supported by the “Strategic Priority Research Program” of the Chinese Academy of Sciences (Grant No. XDB38020400), the CAS Project for Young Scientists in Basic Research (Grant No. YSBR-077), Shanghai Science and Technology Commission Excellent Academic Leaders Program (22XD1424700) and Shanghai Municipal Science and Technology Major Project (Grant No.2017SHZDZX01). XG is also supported by Shanghai Municipal Science and Technology Major Project (Grant No. 2017SHZDZX01). BGF is a National Institute of Health and Care Research Academic Clinical Lecturer and was previously supported by a Medical Research Council (MRC) clinical research training fellowship (MR/ S021280/1). CL is supported by a Sir Henry Dale Fellowship jointly funded by the Wellcome Trust and the Royal Society (223267/Z/21/Z). RE was supported by a Wellcome Trust collaborative award (209233). This research was funded in whole, or in part, by the Wellcome Trust [Grant numbers: 209233, 223267/Z/21/Z]. For the purpose of open access, the author has applied a CC BY public copyright licence to any Author Accepted Manuscript version arising from this submission.

## Author contributions

Conception and study design: JZ, JG, BGF, JHT, XG, SW

Data acquisition: JZ, JG, HL, LJ, XG, SW

Data analysis: JZ, JG

Interpretation of results: JZ, JG, BGF, HL, RE, CL, TC, LJ, JHT, XG, SW

Article drafts were written by JZ, and critically revised by all authors. The final version of the article was approved by all authors.

## References

1. Hunter DJ, Bierma-Zeinstra S. Osteoarthritis. Lancet. 2019;393(10182):1745–59.

2. Zheng J, Frysz M, Faber BG, Lin H, Ebsim R, Ge J, et al. Comparison between UK Biobank and Shanghai Changfeng suggests distinct hip morphology may contribute to ethnic differences in the prevalence of hip osteoarthritis. Osteoarthritis and Cartilage. 2023.

3. Lau EM, Lin F, Lam D, Silman A, Croft P. Hip osteoarthritis and dysplasia in Chinese men. Ann Rheum Dis. 1995;54(12):965–9.

4. Iidaka T, Muraki S, Akune T, Oka H, Kodama R, Tanaka S, et al. Prevalence of radiographic hip osteoarthritis and its association with hip pain in Japanese men and women: the ROAD study. Osteoarthritis Cartilage. 2016;24(1):117–23.

5. Park JH, Lee JS, Lee SJ, Kim YH. Low prevalence of radiographic hip osteoarthritis and its discordance with hip pain: A nationwide study in Korea. Geriatr Gerontol Int. 2021;21(1):20–6.

6. Faber BG, Bredbenner TL, Baird D, Gregory J, Saunders F, Giuraniuc CV, et al. Subregional statistical shape modelling identifies lesser trochanter size as a possible risk factor for radiographic hip osteoarthritis, a cross-sectional analysis from the Osteoporotic Fractures in Men Study. Osteoarthritis Cartilage. 2020;28(8):1071–8.

7. Faber BG, Baird D, Gregson CL, Gregory JS, Barr RJ, Aspden RM, et al. DXA-derived hip shape is related to osteoarthritis: findings from in the MrOS cohort. Osteoarthritis Cartilage. 2017;25(12):2031–8.

8. Baird DA, Evans DS, Kamanu FK, Gregory JS, Saunders FR, Giuraniuc CV, et al. Identification of Novel Loci Associated With Hip Shape: A Meta-Analysis of Genomewide Association Studies. J Bone Miner Res. 2019;34(2):241–51.

9. Faber BG, Frysz M, Hartley AE, Ebsim R, Boer CG, Saunders FR, et al. A Genome-Wide Association Study Meta-Analysis of Alpha Angle Suggests Cam-Type Morphology May Be a Specific Feature of Hip Osteoarthritis in Older Adults. Arthritis & Rheumatology. 2023;75(6):900–9.

10. Faber BG, Frysz M, Boer CG, Evans DS, Ebsim R, Flynn KA, et al. The identification of distinct protective and susceptibility mechanisms for hip osteoarthritis: findings from a genome-wide association study meta-analysis of minimum joint space width and Mendelian randomisation cluster analyses. eBioMedicine. 2023;95.

11. Gao X, Hofman A, Hu Y, Lin H, Zhu C, Jeekel J, et al. The Shanghai Changfeng Study: a community-based prospective cohort study of chronic diseases among middle-aged and elderly: objectives and design. Eur J Epidemiol. 2010;25(12):885–93.

12. Frysz M, Faber BG, Ebsim R, Saunders FR, Lindner C, Gregory JS, et al. Machine-learning derived acetabular dysplasia and cam morphology are features of severe hip osteoarthritis: findings from UK Biobank. J Bone Miner Res. 2022.

13. Heppenstall SV, Ebsim R, Saunders FR, Lindner C, Gregory JS, Aspden RM, et al. Hip geometric parameters are associated with radiographic and clinical hip osteoarthritis: findings from a cross-sectional study in UK Biobank. medRxiv preprint. 2023.

14. Faber B. Geometric Parameters Python 3.0 Code. GitHub 2022 [Available from: https://zenodo.org/badge/latestdoi/518486087.

15. Zeng H, Ge J, Xu W, Ma H, Chen L, Xia M, et al. Twelve Loci Associated With Bone Density in Middle-aged and Elderly Chinese: The Shanghai Changfeng Study. The Journal of Clinical Endocrinology & Metabolism. 2023;108(2):295–305.

16. Purcell S, Neale B, Todd-Brown K, Thomas L, Ferreira MAR, Bender D, et al. PLINK: A Tool Set for Whole-Genome Association and Population-Based Linkage Analyses. The American Journal of Human Genetics. 2007;81(3):559–75.

17. Delaneau O, Coulonges C, Zagury J-F. Shape-IT: new rapid and accurate algorithm for haplotype inference. BMC Bioinformatics. 2008;9(1).

18. Schork NJ, Howie BN, Donnelly P, Marchini J. A Flexible and Accurate Genotype Imputation Method for the Next Generation of Genome-Wide Association Studies. PLoS Genetics. 2009;5(6).

19. Heppenstall SV, Ebsim R, Saunders FR, Lindner C, Gregory JS, Aspden RM, et al. Hip geometric parameters are associated with radiographic and clinical hip osteoarthritis: Findings from a cross-sectional study in UK Biobank. Osteoarthritis Cartilage. 2023.

20. Jiang L, Zheng Z, Qi T, Kemper KE, Wray NR, Visscher PM, et al. A resource-efficient tool for mixed model association analysis of large-scale data. Nature Genetics. 2019;51(12):1749–55.

21. D. Turner S. qqman: an R package for visualizing GWAS results using Q-Q and manhattan plots. Journal of Open Source Software. 2018;3(25):731.

22. Yang J, Lee SH, Goddard ME, Visscher PM. GCTA: A Tool for Genome-wide Complex Trait Analysis. The American Journal of Human Genetics. 2011;88(1):76–82.

23. Yang J, Ferreira T, Morris AP, Medland SE, Madden PAF, Heath AC, et al. Conditional and joint multiple-SNP analysis of GWAS summary statistics identifies additional variants influencing complex traits. Nature Genetics. 2012;44(4):369–75.

24. Pruim RJ, Welch RP, Sanna S, Teslovich TM, Chines PS, Gliedt TP, et al. LocusZoom: regional visualization of genome-wide association scan results. Bioinformatics. 2010;26(18):2336–7.

25. Lee SH, Yang J, Goddard ME, Visscher PM, Wray NR. Estimation of pleiotropy between complex diseases using single-nucleotide polymorphism-derived genomic relationships and restricted maximum likelihood. Bioinformatics. 2012;28(19):2540–2.

26. Styrkarsdottir U, Stefansson OA, Gunnarsdottir K, Thorleifsson G, Lund SH, Stefansdottir L, et al. GWAS of bone size yields twelve loci that also affect height, BMD, osteoarthritis or fractures. Nat Commun. 2019;10(1):2054.

27. Zheng HF, Forgetta V, Hsu YH, Estrada K, Rosello-Diez A, Leo PJ, et al. Whole-genome sequencing identifies EN1 as a determinant of bone density and fracture. Nature. 2015;526(7571):112-7.

28. Nethander M, Coward E, Reimann E, Grahnemo L, Gabrielsen ME, Wibom C, et al. Assessment of the genetic and clinical determinants of hip fracture risk: Genome-wide association and Mendelian randomization study. Cell Reports Medicine. 2022;3(10).

29. Boer CG, Hatzikotoulas K, Southam L, Stefánsdóttir L, Zhang Y, Coutinho de Almeida R, et al. Deciphering osteoarthritis genetics across 826,690 individuals from 9 populations. Cell. 2021;184(18):4784–818.e17.

30. Watanabe K, Taskesen E, van Bochoven A, Posthuma D. Functional mapping and annotation of genetic associations with FUMA. Nature Communications. 2017;8(1).

31. Kircher M, Witten DM, Jain P, O’Roak BJ, Cooper GM, Shendure J. A general framework for estimating the relative pathogenicity of human genetic variants. Nat Genet. 2014;46(3):310–5.

32. Boyle AP, Hong EL, Hariharan M, Cheng Y, Schaub MA, Kasowski M, et al. Annotation of functional variation in personal genomes using RegulomeDB. Genome Research. 2012;22(9):1790–7.

33. Ernst J, Kellis M. ChromHMM: automating chromatin-state discovery and characterization. Nature Methods. 2012;9(3):215–6.

34. Lonsdale J, Thomas J, Salvatore M, Phillips R, Lo E, Shad S, et al. The Genotype-Tissue Expression (GTEx) project. Nature Genetics. 2013;45(6):580–5.

35. Keen J, Moore H. The Genotype-Tissue Expression (GTEx) Project: Linking Clinical Data with Molecular Analysis to Advance Personalized Medicine. Journal of Personalized Medicine. 2015;5(1):22–9.

36. Carithers LJ, Moore HM. The Genotype-Tissue Expression (GTEx) Project. Biopreservation and Biobanking. 2015;13(5):307–8.

37. Baird DA, Paternoster L, Gregory JS, Faber BG, Saunders FR, Giuraniuc CV, et al. Investigation of the Relationship Between Susceptibility Loci for Hip Osteoarthritis and Dual X- Ray Absorptiometry–Derived Hip Shape in a Population - Based Cohort of Perimenopausal Women. Arthritis &amp; Rheumatology. 2018;70(12):1984–93.

38. Abdulrahim H, Jiao Q, Swain S, Sehat K, Sarmanova A, Muir K, et al. Constitutional morphological features and risk of hip osteoarthritis: a case-control study using standard radiographs. Ann Rheum Dis. 2021;80(4):494–501.

39. van Buuren MMA, Arden NK, Bierma-Zeinstra SMA, Bramer WM, Casartelli NC, Felson DT, et al. Statistical shape modeling of the hip and the association with hip osteoarthritis: a systematic review. Osteoarthritis Cartilage. 2021;29(5):607–18.

40. Pineault KM, Wellik DM. Hox Genes and Limb Musculoskeletal Development. Current Osteoporosis Reports. 2014;12(4):420–7.

41. Morris JA, Kemp JP, Youlten SE, Laurent L, Logan JG, Chai RC, et al. An atlas of genetic influences on osteoporosis in humans and mice. Nature Genetics. 2018;51(2):258–66.

42. Yengo L, Vedantam S, Marouli E, Sidorenko J, Bartell E, Sakaue S, et al. A saturated map of common genetic variants associated with human height. Nature. 2022;610(7933):704–12.

43. Gkiatas I, Boptsi A, Tserga D, Gelalis I, Kosmas D, Pakos E. Developmental dysplasia of the hip: a systematic literature review of the genes related with its occurrence. EFORT Open Reviews. 2019;4(10):595–601.

44. Miyamoto Y, Mabuchi A, Shi D, Kubo T, Takatori Y, Saito S, et al. A functional polymorphism in the 5′ UTR of GDF5 is associated with susceptibility to osteoarthritis. Nature Genetics. 2007;39(4):529–33.

45. Dai J, Shi D, Zhu P, Qin J, Ni H, Xu Y, et al. Association of a single nucleotide polymorphism in growth differentiate factor 5 with congenital dysplasia of the hip: a case-control study. Arthritis Research & Therapy. 2008;10(5).

46. Vaes RBA, Rivadeneira F, Kerkhof JM, Hofman A, Pols HAP, Uitterlinden AG, et al. Genetic variation in the GDF5 region is associated with osteoarthritis, height, hip axis length and fracture risk: the Rotterdam study. Annals of the Rheumatic Diseases. 2008;68(11):1754–60.

47. Mailund T, Wu D-D, Li G-M, Jin W, Li Y, Zhang Y-P. Positive Selection on the Osteoarthritis-Risk and Decreased-Height Associated Variants at the GDF5 Gene in East Asians. PLoS ONE. 2012;7(8).

48. Huang Q, Sun Y, Wang C, Hao Z, Dai J, Chen D, et al. A Common Variant Of Ubiquinol-Cytochrome c Reductase Complex Is Associated with DDH. Plos One. 2015;10(4).

49. Gumus E, Temiz E, Sarikaya B, Yuksekdag O, Sipahioglu S, Gonel A. The Association Between BMP-2, UQCC1 and CX3CR1 Polymorphisms and the Risk of Developmental Dysplasia of the Hip. Indian Journal of Orthopaedics. 2020;55(1):169–75.

50. Bulik-Sullivan B, Finucane HK, Anttila V, Gusev A, Day FR, Loh PR, et al. An atlas of genetic correlations across human diseases and traits. Nat Genet. 2015;47(11):1236–41.

51. Sanderson E, Glymour MM, Holmes MV, Kang H, Morrison J, Munafò MR, et al. Mendelian randomization. Nature Reviews Methods Primers. 2022;2(1):6.

52. Littlejohns TJ, Holliday J, Gibson LM, Garratt S, Oesingmann N, Alfaro-Almagro F, et al. The UK Biobank imaging enhancement of 100,000 participants:Jrationale, data collection, management and future directions. Nature Communications. 2020;11(1):2624.

53. Kurki MI, Karjalainen J, Palta P, Sipilä TP, Kristiansson K, Donner K, et al. FinnGen: Unique genetic insights from combining isolated population and national health register data. 2022.

